# Circulating tumor DNA association with residual cancer burden after neoadjuvant chemotherapy in triple-negative breast cancer in TBCRC 030

**DOI:** 10.1101/2023.03.06.23286772

**Authors:** Heather A. Parsons, Timothy Blewett, Xiangying Chu, Sainetra Sridhar, Katheryn Santos, Kan Xiong, Vandana G. Abramson, Ashka Patel, Ju Cheng, Adam Brufsky, Justin Rhoades, Jeremy Force, Ruolin Liu, Tiffany A. Traina, Lisa A. Carey, Mothaffar F. Rimawi, Kathy D. Miller, Vered Stearns, Jennifer Specht, Carla Falkson, Harold J. Burstein, Antonio C. Wolff, Eric P. Winer, Nabihah Tayob, Ian E. Krop, G. Mike Makrigiorgos, Todd R. Golub, Erica L. Mayer, Viktor A. Adalsteinsson

**Affiliations:** Medical Oncology, Dana-Farber Cancer Institute, Boston, MA, USA; Breast Oncology Program, Dana-Farber Brigham Cancer Center, Boston, MA, USA; Harvard Medical School, Boston, MA, USA; Broad Institute of MIT and Harvard, Boston, MA, USA; Data Science, Dana-Farber Cancer Institute, Boston, MA, USA; Vanderbilt-Ingram Cancer Center, Nashville, TN, USA; University of Pittsburgh School of Medicine, Pittsburgh, PA, USA; Duke Cancer Center, Durham, NC, USA; Memorial Sloan Kettering Cancer Center, New York, NY, USA; The University of North Carolina Lineberger Comprehensive Cancer Center, Chapel Hill, NC, USA; Baylor College of Medicine Dan L. Duncan Comprehensive Cancer Center, Houston, TX, USA; Indiana University Melvin and Bren Simon Comprehensive Cancer Center, Indianapolis, IN, USA; Birmingham, AB, USA; Johns Hopkins Sidney Kimmel Comprehensive Cancer Center, Baltimore, MD, USA; Seattle Cancer Care Alliance, Seattle, WA, USA; The University of Alabama at Birmingham, Birmingham, AL, USA; Radiation Oncology, Dana-Farber Cancer Institute, Boston, MA, USA

**Keywords:** circulating tumor DNA, triple-negative breast cancer, biomarkers, recurrence, minimal residual disease

## Abstract

**Purpose:** To examine circulating tumor DNA (ctDNA) and its association with residual cancer burden (RCB) using an ultrasensitive assay in patients with triple-negative breast cancer (TNBC) receiving neoadjuvant chemotherapy (NAT).

**Patients and Methods:** We identified responders (RCB-0/1) and matched non-responders (RCB-2/3) from the phase II TBCRC 030 prospective study of neoadjuvant paclitaxel vs. cisplatin in TNBC. We collected plasma samples at baseline, three weeks, and twelve weeks (end of therapy). We created personalized ctDNA assays utilizing MAESTRO mutation enrichment sequencing. We explored associations between ctDNA and RCB status and disease recurrence.

**Results:** Of 139 patients, 68 had complete samples and no additional NAT. Twenty-two were responders and 19 of those had sufficient tissue for whole-genome sequencing. We identified an additional 19 non-responders for a matched case-control analysis of 38 patients using a MAESTRO ctDNA assay tracking 319-1000 variants (median 1000) to 114 plasma samples from 3 timepoints. Overall, ctDNA positivity was 100% at baseline, 79% at week 3, and 55% at week 12. Median tumor fraction (TFx) was 3.7 × 10^−4^ (range: 7.9 × 10^−7^ to 4.9 × 10^−1^). TFx decreased 285-fold from baseline to week 3 in responders and 24-fold in non-responders. Week 12 ctDNA clearance correlated with RCB: clearance was observed in 10/11 patients with RCB-0, 3/8 with RCB-1, 4/15 with RCB-2, and 0/4 with RCB-3. Among 6 patients with known recurrence five had persistent ctDNA at week 12.

**Conclusion:** NAT for TNBC reduced ctDNA TFx by 285-fold in responders and 24-fold in non-responders. In 58% (22/38) of patients, ctDNA TFx dropped below the detection level of a commercially available test, emphasizing the need for sensitive tests. Additional studies will determine if ctDNA-guided approaches can improve outcomes.

## Introduction

Triple-negative breast cancer (TNBC) comprises 15% to 20% of invasive breast cancer cases.^1^ Compared to patients with other breast cancer subtypes, those with early-stage TNBC (eTNBC) face a higher risk of distant recurrence and death within 3-5 years of diagnosis.^2-4^ Patients who experience a pathologic complete response (pCR) to neoadjuvant systemic therapy have significantly lower rates of recurrence and improved survival outcomes compared to patients who have residual invasive disease in the breast or lymph nodes.^5-7^ The presence or absence of pCR and the quantitative measure of residual cancer burden (RCB) are prognostic markers and may guide adjuvant systemic therapy after neoadjuvant treatment for eTNBC.^8,9^

Isolation of circulating tumor DNA (ctDNA) from cell-free DNA (cfDNA) in the plasma offers a minimally invasive means of evaluating disease status and response to treatment in real time in patients with early-stage breast cancer. Persistence of ctDNA after neoadjuvant therapy is associated with higher risk of distant recurrence.^10-12^ Despite strong positive predictive value in some studies, ctDNA levels (as measured by commercially available assays^13^) can become undetectable for most patients who still have residual disease, suggesting the need for more sensitive assays to detect circulating minimal residual disease (MRD) to potentially guide optimization of neoadjuvant and adjuvant therapy.^11,14^

TBCRC 030 (NCT01982448) was a prospective, randomized phase II study of 12 weeks of neoadjuvant cisplatin or paclitaxel for patients with TNBC. The primary objective was to evaluate the utility of a homologous recombination deficiency (HRD) biomarker for predicting RCB status by treatment arm.^15^ In this exploratory analysis, we evaluated ctDNA dynamics in patients who responded (RCB 0/1) or did not respond (RCB 2/3) to neoadjuvant chemotherapy. We reasoned that a more sensitive ctDNA approach would enable improved resolution of responses to therapy. We applied a recently developed, tumor-informed, mutation enrichment sequencing assay called MAESTRO^16^ to track up to one thousand patient-specific tumor mutations in cfDNA with a reported sensitivity that is 100-fold higher than existing, commercial ctDNA tests and explore associations between levels of detectable ctDNA and pathologic response.

## Methods

### Study Population

TBCRC 030 was an investigator-initiated, prospective, open-label, randomized phase II trial that enrolled patients with invasive TNBC or estrogen receptor [ER]-low expressing breast cancer (estrogen receptor [ER] ≤ 5%, progesterone receptor [PR] ≤ 5%, HER2 immunohistochemistry [IHC] 0/1+ or fluorescence in situ hybridization ratio [FISH] ratio < 2.0) that was clinical stage I (T1 ≥ 1.5 cm) or stage II-III.^15^ Patients with a known germline *BRCA1/2* mutation were not eligible, although baseline genetic testing was not mandated. Patients were randomly assigned in a 1:1 ratio to receive either cisplatin 75 mg/m^2^ every three weeks for four cycles or weekly paclitaxel 80 mg/m^2^ for 12 weeks. After the completion of neoadjuvant chemotherapy, patients could proceed to definitive breast surgery. The primary study endpoint was pathologic response assessed using the RCB score.^17^ Patients with RCB 0/1 were considered responders, whereas patients with RCB 2/3 were considered non-responders. Patients who were not ready for surgery after the initial 12 weeks of study therapy were allowed to cross over to an alternative additional neoadjuvant chemotherapy regimen and were considered non-responders (RCB 2/3). The primary objective of the parent study was to correlate baseline biomarker for HRD and RCB by study arm. This study was approved by the Dana-Farber/Harvard Cancer Center Institutional Review Board. All patients provided informed consent. This study was conducted in compliance with the principles of the Declaration of Helsinki.

For this exploratory analysis, responders (RCB 0/1) and non-responders (RCB 2/3) from both study arms who did not receive additional neoadjuvant therapy prior to surgery were selected for analysis from the study cohort, matched on baseline nodal status and tumor size. As a post hoc study amendment, available patients were followed for event-free survival (EFS).

### ctDNA Analysis

Serial blood samples for ctDNA analysis were collected prior to treatment initiation (W0), at three weeks of therapy (W3), and at 12 weeks of therapy (W12), prior to surgery. Whole-genome sequencing (WGS) was performed on primary tumor tissue from the baseline biopsy and matched normal genomic DNA from the baseline whole blood sample to identify somatic mutations. These somatic mutations were used to design two tumor-informed ctDNA assays tracking up to 1000 mutations for each patient. The first assay, MRD Tracker, was an optimized version of the Parsons et al,^18^ tumor-informed, hybrid-capture duplex sequencing MRD test; and the second assay, MAESTRO, was a tumor-informed, mutation enrichment sequencing MRD test.^16^ MAESTRO improves on MRD Tracker by incorporating mutation enrichment.^16^ Both assays were applied to all patient-specific plasma, tumor tissue and matched normal samples. After consensus calling and filtering, we subsetted to mutations that were validated in the tumor tissue and not detected in the matched normal to ensure we were tracking true somatic mutations. Additionally, plasma samples with 10 or fewer mutations detected were flagged for semi-automated review prior to unblinding us to the response status. In these cases, each detected mutation was manually reviewed to determine if the mutation was likely false, in which case it was discarded from all patient samples. Finally, MRD status, tumor fraction (TFx) and limit of detection (LOD) – the TFx at which detection power is 90% - were generated for both assays as described previously.^18^ We computed the TFx fold change for W3 and W12 samples with respect to W0 by substituting the LOD for TFx when the sample was MRD negative.

## Results

### Study Design and ctDNA Testing

Among 139 study participants, 68 had complete tumor tissue and plasma samples, and no receipt of crossover neoadjuvant therapy after 12 weeks of study therapy. Twenty-two of these 68 (32%) responded to neoadjuvant chemotherapy (RBC 0/1) and 19 had sufficient tumor mass and quality to yield high quality whole genome sequencing data. These 19 responders were analyzed along with 19 matched non-responders (RCB 2/3, **Figure 1A**).

**Figure 1:**
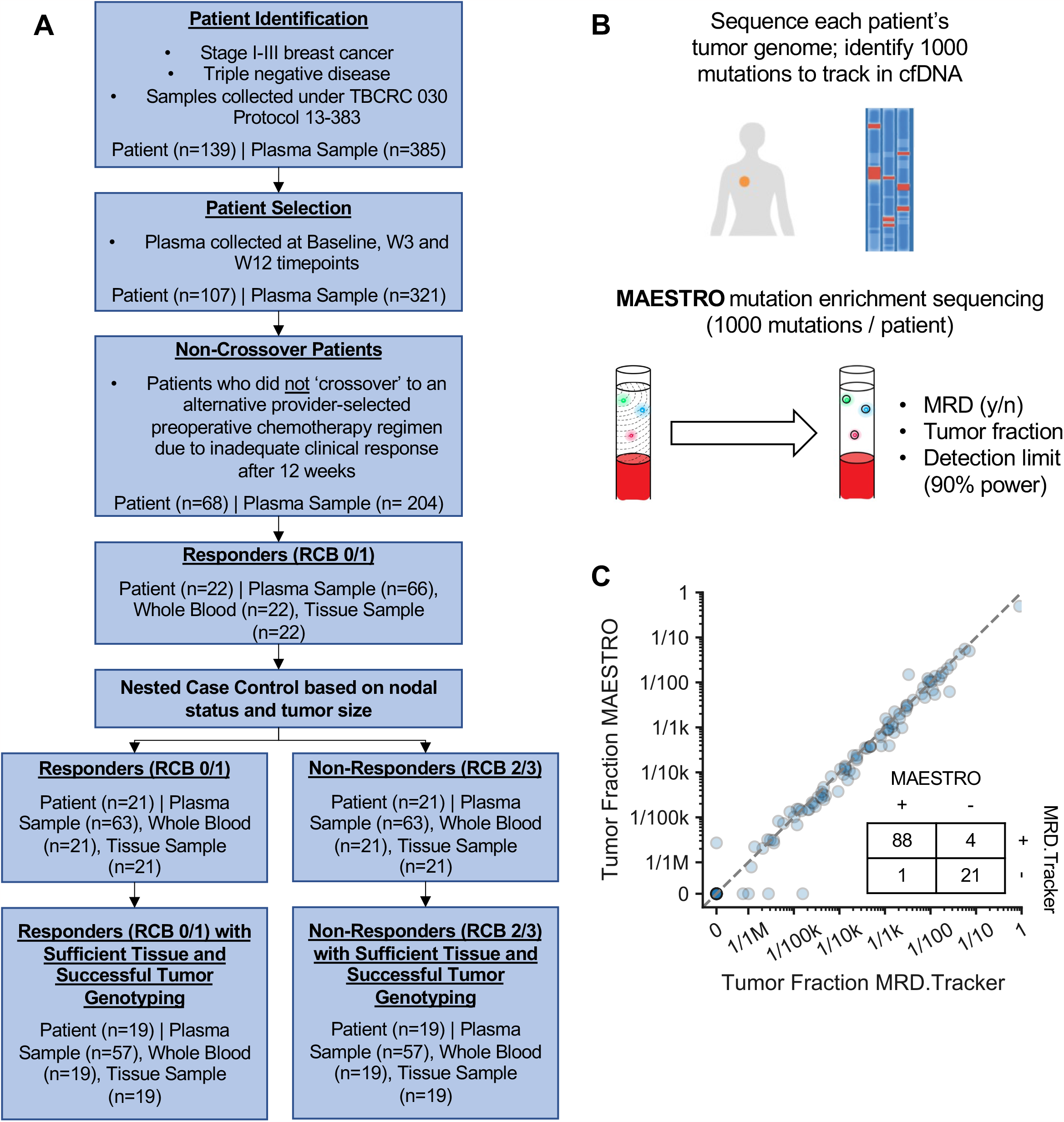
REMARK diagram and MAESTRO ctDNA assay. Depicts (A) how the cohort of patients were selected from the 13-383 study population and organized into case control pairs and (B) how bespoke MAESTRO assays were designed for ctDNA detection. (C) MAESTRO results were validated against MRD.Tracker – a complementary MRD assay without mutation enrichment. The number of ctDNA positive and negative samples using each assay is denoted in the inset table. Abbreviations: ctDNA, circulating tumor DNA; MRD, minimal residual disease; RCB, residual cancer burden; W, week

Personalized MAESTRO tumor-informed, mutation enrichment sequencing MRD assays were designed targeting 319-1000 (median 1000) genome-wide, somatic mutations per patient and applied to 114 plasma samples (W0 = 38; W3 = 38; W12 = 38, **Figure 1B**, 13). For comparison, personalized MRD Tracker assays, optimized from our previously validated Parsons et al method,^18^ were also designed targeting 434-1000 (median 1000) genome-wide, somatic mutations per patient and applied to all the same plasma samples. This served to confirm concordant results between MAESTRO and an orthogonally validated test.

For each patient, a median of 633 mutations (range 269-686), or 48% (range 45%-58%) of all tracked mutations, was tracked using both assays. For mutations detected by both, MAESTRO showed strong enrichment in variant allele frequencies (**Supplementary Fig 1**). Despite differences in mutations tracked and methodologies used, MAESTRO yielded highly concordant ctDNA tumor fractions down to low parts-per-million (r^2=0.997, **Figure 1C**). Of the 5 samples with discordant results, all 5 had TFx lower than estimated detection limits at 90% power, consistent with less reliable detection (**Supplementary Fig 2**). Our results demonstrate that MAESTRO enriches rare mutations and detects MRD at similar ctDNA TFx as an orthogonally validated test.

### ctDNA Dynamics

We next sought to examine how ctDNA dynamics from MAESTRO testing associated with response to preoperative chemotherapy. At W0, all 38 patients (100%) were positive for ctDNA, with a median TFx of 3.1 × 10^−3^ (range: 2.2 × 10^−6^ to 4.9 × 10^−1^). Tumor stage and nodal status were associated with TFx. In patients with T1-T2 tumors, the median TFx was 2.7 × 10^−3^ (range: 2.2 × 10^−6^ to 5.5 × 10^−2^), while median TFx was 2.7 × 10^−1^ (range: 4.3 × 10^−2^ to 4.9 × 10^−1^) in patients with T3-T4 tumors. Additionally, patients with positive axillary lymph nodes had a median TFx of 4.7 × 10^−3^ (range 2.4 × 10^−4^ to 5.5 × 10^−2^) while patients with negative nodes had 1.4 × 10^−3^ (range 2.2 × 10^−6^ to 4.9 × 10^−1^). At W3 and W12, n = 30 patients (79%) and n = 21 patients (55%) were positive for ctDNA, respectively (**Figure 2A, Supplementary Fig 3, Supplementary Fig 4**).

**Figure 2:**
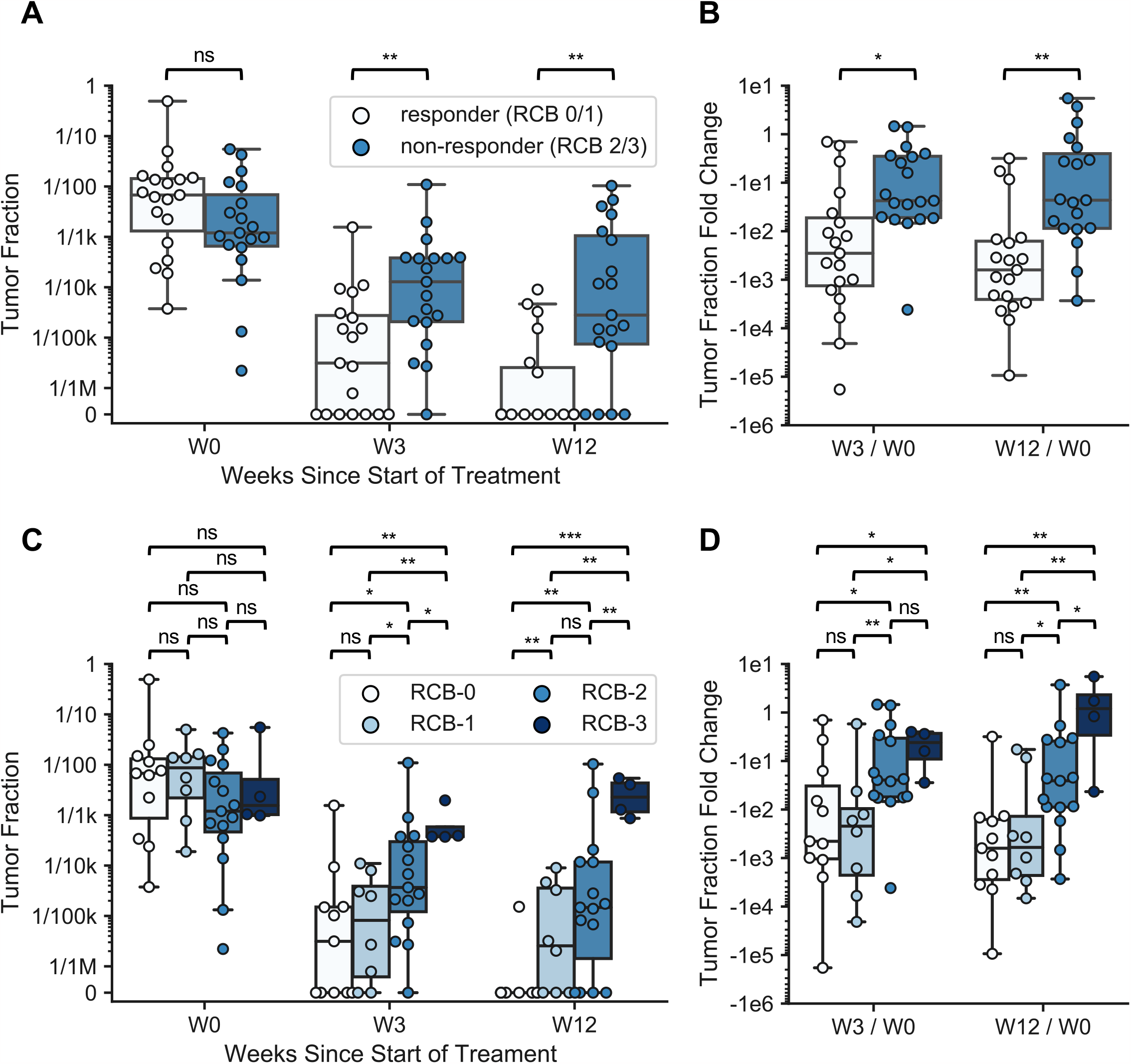
ctDNA dynamics correlated with response status and RCB score. TFxs observed during neoadjuvant therapy separated by (A) response status and (C) RCB score. Additionally, TFx fold change relative to baseline observed separated by (B) response status and (D) RCB score. Statistical significance was evaluated using the Wilcoxon signed-rank test (A & B) and the Mann-Whitney U test (C & D) (*: p < 0.05; **: p < 0.01; ***: p < 0.001). Abbreviations: ctDNA, circulating tumor DNA; RCB, residual cancer burden; TFx, tumor fraction; W, week

Among responders (n = 19), the TFx decreased 285-fold from W0 to W3. By contrast, the TFx decreased 24-fold from W0 to W3 among non-responders (**Table 1, Figure 2B**). At W12, ctDNA was cleared in 10 of 11 patients with RCB 0, 3 of 8 patients with RCB 1, 4 of 15 patients with RCB 2, and 0 of 4 patients with RCB 3 (**Figure 2C**). Analysis of TFx within cfDNA was more specific than ultrasound at detecting MRD (**Figure 3**). Pathology analysis also had higher concordance with ctDNA than with imaging when residual disease burden was low.

**Table 1:** Tumor fraction (TFx) and tumor fraction fold change by response to neoadjuvant therapy.

|  | Responders<br>(n=19) | Non-Responders<br>(n=19) |
| --- | --- | --- |
| **TFx** |  |  |
| **Median (Min, Max)** |  |  |
| W0 | $6.8 \times 10^{-3}$ ($3.7 \times 10^{-5}$, $4.9 \times 10^{-1}$) | $1.2 \times 10^{-3}$ ($2.2 \times 10^{-6}$, $5.5 \times 10^{-2}$) |
| W3 | $3.1 \times 10^{-6}$ (0, $1.6 \times 10^{-3}$) | $1.3 \times 10^{-4}$ (0, $1.1 \times 10^{-2}$) |
| W12 | 0 (0, $9.0 \times 10^{-5}$) | $2.8 \times 10^{-5}$ (0, $1.0 \times 10^{-2}$) |
| **TFx Fold Change** |  |  |
| **Median (Min, Max)** |  |  |
| From W0 to W3 | $3.5 \times 10^{-3}$ ($5.5 \times 10^{-6}$, $7.0 \times 10^{-1}$) | $4.3 \times 10^{-2}$ ($2.4 \times 10^{-4}$, $1.5 \times 10^0$) |
| From W0 to W12 | $1.4 \times 10^{-3}$ ($1.1 \times 10^{-5}$, $3.2 \times 10^{-1}$) | $4.4 \times 10^{-2}$ ($3.7 \times 10^{-4}$, $5.5 \times 10^0$) |

**Figure 3:**
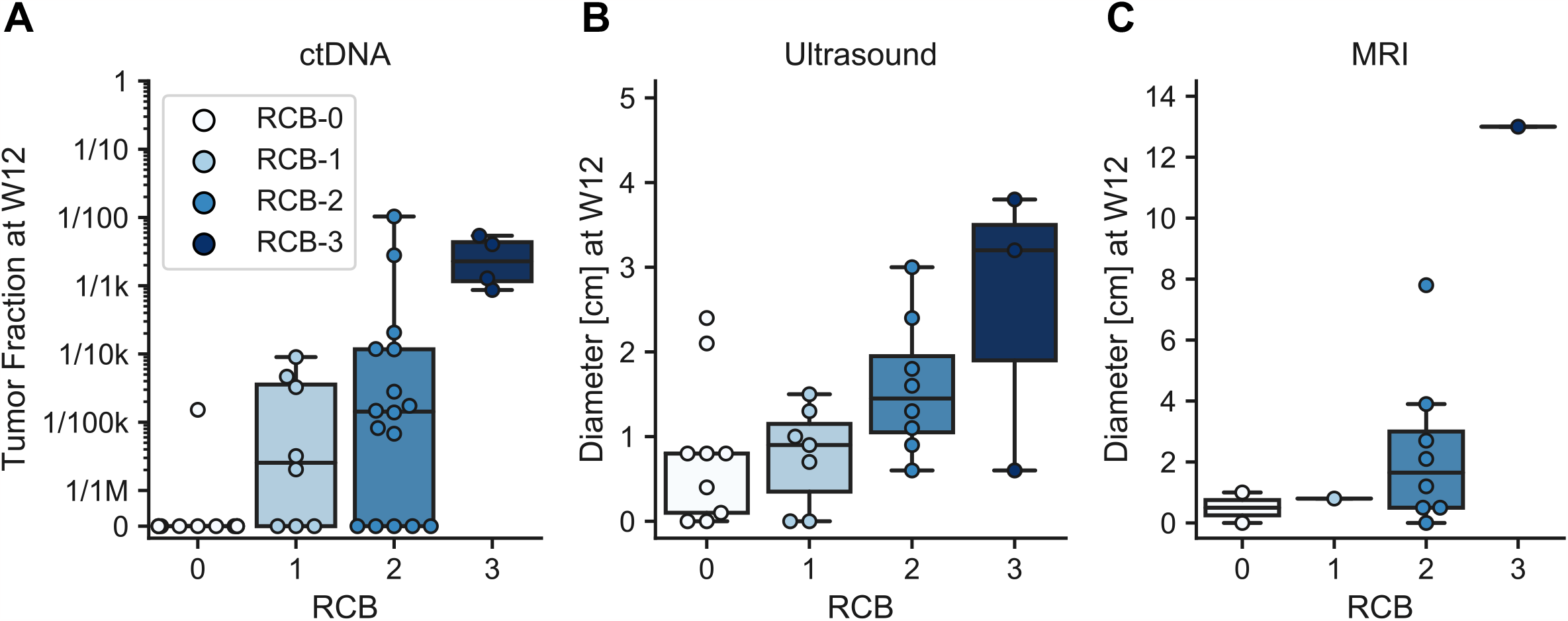
ctDNA TFx prior to surgery correlated with RCB score relative to imaging. Patients RCB score at time of surgery compared against (A) ctDNA TFx, (B) ultrasound diameter, (C) and MRI diameter after completion of neoadjuvant therapy. Abbreviations: ctDNA, circulating tumor DNA; RCB, residual cancer burden; TFx, tumor fraction; W, week

As an additional exploratory analysis, we also simulated the results that we would have obtained had the detection limit of our assay been less sensitive, e.g., only 0.01%, or %, as per many existing tests. At 0.1%, ctDNA detection would have been negative in 34% (13/38), 90% (27/30) and 76% (16/21) of ctDNA positive samples at baseline, W3 and W12, respectively (**Supplementary Fig 5A; Supplementary Data**). Similarly, at 0.01%, ctDNA detection would have been negative in 8% (3/38), 60% (18/30) and 57% (12/21) of ctDNA positive samples at baseline, W3 and W12, respectively. Our findings suggest that the ultrasensitive assay is needed to predict therapy response.

### ctDNA and Disease Recurrence

To investigate whether ctDNA persistence after neoadjuvant therapy was associated with disease recurrence, we analyzed a separate group of 8 patients with known recurrence and 8 without known recurrence (**Figure 4**). Among the 8 patients with documented recurrence, 6 had plasma collected at W12 and 5/6 of these had persistent ctDNA with a median TFx of 4.0 × 10^−3^ (range: 2.0 × 10^−5^ to 7.9 × 10^−2^). The one patient without persistent ctDNA at W12 had only 1 mutation detected, suggesting that its TFx might be below the observed LOD of 1.5 × 10^−5^. Among the 8 patients without known recurrence, 7 had plasma collected at W12 and 5/7 had cleared ctDNA. The 2 patients with persistent ctDNA at W12 had TFxs of 1.5 × 10^−5^ and 2.9 × 10^−5^, which was less than 6/6 and 5/6 patients with known recurrence at W12, respectively. Moreover, all 8 patients without known recurrence had RCB 0, whereas the patients with documented recurrence had RCB 1 (n = 2), RCB 2 (n = 2), or RCB 3 (n = 4).

**Figure 4:**
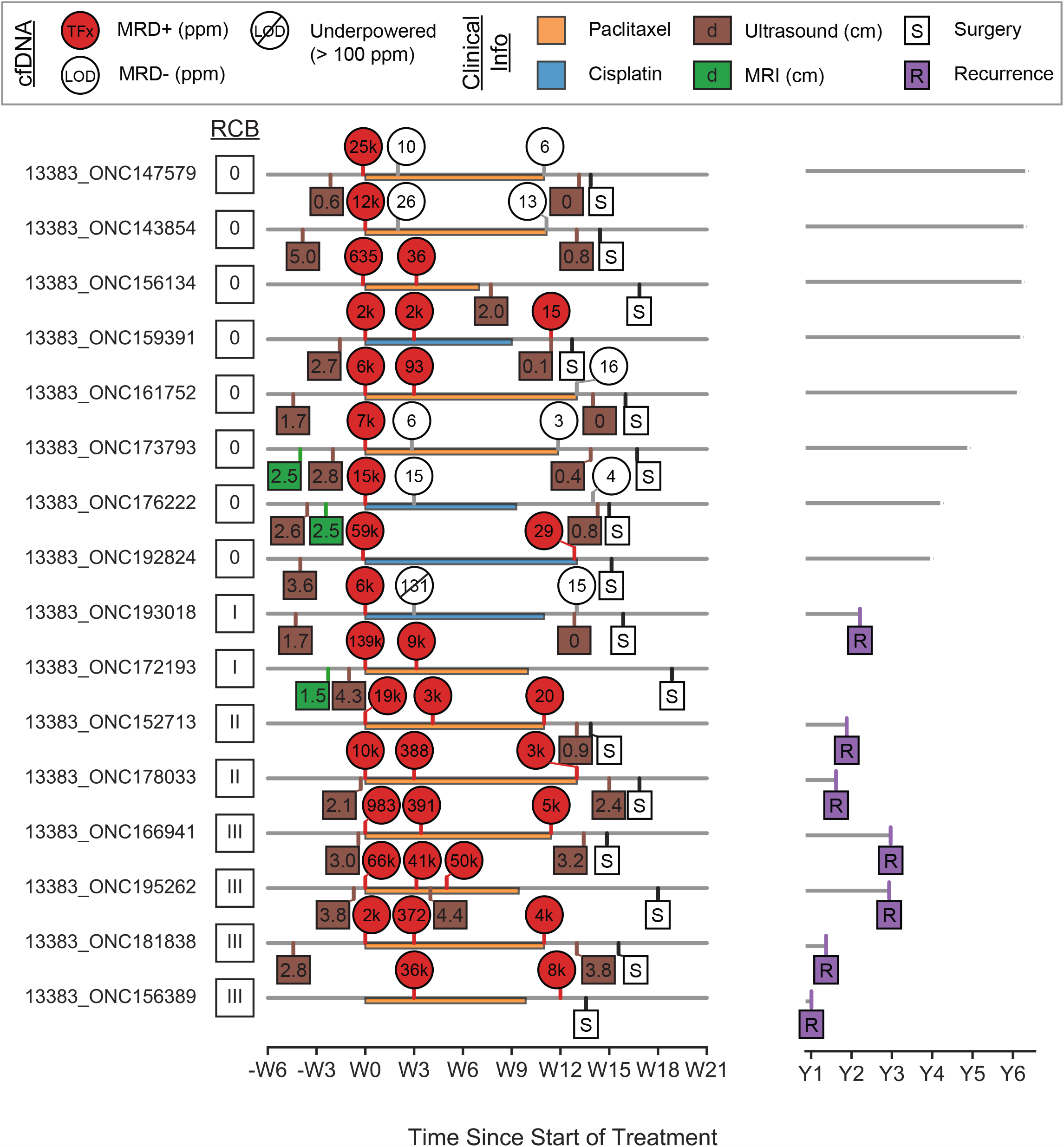
Patients with known distant recurrence had detectable ctDNA prior to surgery unlike most patients without known distant recurrence. 16 patients – 8 with known distant recurrence and 8 without – were selected to analyze whether ctDNA presence after neoadjuvant therapy was associated with distant recurrence. Samples with LODs > 1 × 10^−4^ were considered underpowered, likely due to technical issues. Abbreviations: ctDNA, circulating tumor DNA; LOD, limit of detection; MRD, minimal residual disease; PPM, parts per million; RCB, residual cancer burden; W, week; Y, year

## Discussion

We present a nested case-control study to evaluate ctDNA prevalence and dynamics among 38 patients (19 responders and 19 non-responders) with stage II-III TNBC treated with neoadjuvant cisplatin or paclitaxel. We evaluated the association between RCB and ctDNA detection using the ultrasensitive ctDNA enrichment MAESTRO assay, showing a strong relationship after the completion of neoadjuvant therapy. In a post-hoc, exploratory subset of patients, we detected ctDNA at W12 in 5/6 patients with known distant recurrence and in 2/7 without recurrence. All patients received postoperative chemotherapy but blood samples were not available after completion of treatment. We found that our use of a highly sensitive ctDNA assay was required in order to detect ctDNA at baseline and monitor response at W3 and W12.

Our study is limited by the lack of blood sampling in the post-operative and post-therapeutic settings. Historically, preoperative studies have not collected postoperative biospecimens or patient outcomes, and here our EFS analysis was done post-hoc. Though inclusion of these data adds expense to trials, we encourage ongoing and future neoadjuvant studies to collect them to enable important correlative analyses. Our study is also limited by the small number of patients in the responder and non-responder groups. The application of this method to larger patient groups will allow for further exploration of the association of ctDNA with disease recurrence in early-stage TNBC patients with residual invasive disease after the completion of neoadjuvant therapy.

ctDNA analysis is emerging as a valuable prognostic biomarker in breast and other solid tumors. Studies thus far have shown excellent specificity of ctDNA, enabling clinical trials evaluating the impact of additional therapy for ctDNA positive patients (NCT03285412, NCT04567420, NCT04985266, NCT04849364, NCT05512364, NCT05388149). But available tests have lacked sufficient sensitivity to consider truly tailoring therapy, with escalation or change of therapy for patients who remain ctDNA positive after standard treatment and avoidance of additional therapy for ctDNA negative patients. ctDNA dynamics were recently evaluated in 84 patients with high-risk early-stage breast cancer treated with standard neoadjuvant chemotherapy (paclitaxel followed by anthracycline) with or without the AKT inhibitor MK-2206 in the I-SPY2 trial.^11^ Blood was collected for ctDNA analysis at T0 (pre-treatment), T1 (3 weeks after initiation of paclitaxel), T2 (between paclitaxel and anthracycline), and T3 (prior to surgery). At T0, only 61 patients (73%) had detectable ctDNA. Those who remained ctDNA-positive at T1 were significantly less likely to experience a pCR than those who cleared ctDNA (p = 0.012). Among patients who did not experience a pCR, those with detectable ctDNA had a significantly increased risk of metastatic recurrence. However, most patients with residual disease after neoadjuvant therapy had undetectable ctDNA, limiting the actionability of a negative test and emphasizing the utility of more sensitive ctDNA detection.^11^

In conclusion, we conducted a case-control study of the dynamics and association of ctDNA with RCB in patients with eTNBC treated on the TBCRC 030 study of preoperative cisplatin vs. paclitaxel. We used a highly sensitive mutation enrichment approach (MAESTRO) and showed a strong association between ctDNA TFx and RCB. For 58% (22/38) of patients, TFx had dropped below the LOD reported in currently available conventional assays, underscoring the need for more sensitive approaches in this setting to enable treatment tailoring. These data suggest that sensitive ctDNA analysis in the neoadjuvant setting may be a valuable tool to guide treatment in the neoadjuvant setting for eTNBC. If our findings are confirmed, such assays could inform the design of future studies using them as integral biomarkers to tailor therapy for patients with eTNBC.

## Data Availability

The data that support the findings of this study are available from the corresponding authors [HAP, ELM, and VAA], upon reasonable request.

## Acknowledgment of Research Support

The work was supported by NCI Mentored Clinical Scientist Research Career Development Award (1K08CA252639, H.A.P.); NIH (R01 CA221874 to G.M.M.); Gerstner Family Foundation; Forget-Me-Not Fund.

## Acknowledgments

We are grateful to all the patients who generously volunteered to participate in this study. We thank the TBCRC investigators, research nurses, & study coordinators for their efforts on behalf of the patients. We appreciate the funding support provided to the TBCRC by its foundation partners: The Breast Cancer Research Foundation, Susan G. Komen. We thank the Gerstner Family Foundation for its generous support of this work. The authors would like to thank Timothy Erick for medical writing support, as well as Kaitlyn Bifolck for medical editing. Both are full-time employees of Dana-Farber Cancer Institute.

