## Supplemental Figures for "Circulating tumor DNA association with residual cancer burden after neoadjuvant chemotherapy in triple-negative breast cancer in TBCRC 030"

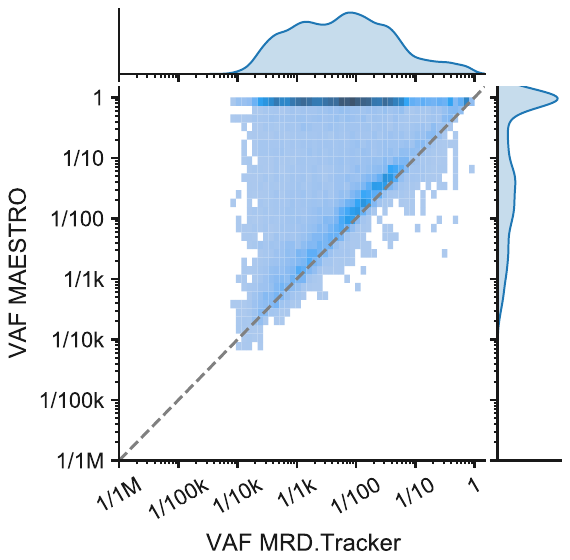

**Supplementary Figure 1: MAESTRO enrichment relative to MRD.Tracker.** MAESTRO enrichment was assessed by comparing the variant allele fraction (VAF) at mutation sites detected with MAESTRO and MRD.Tracker.

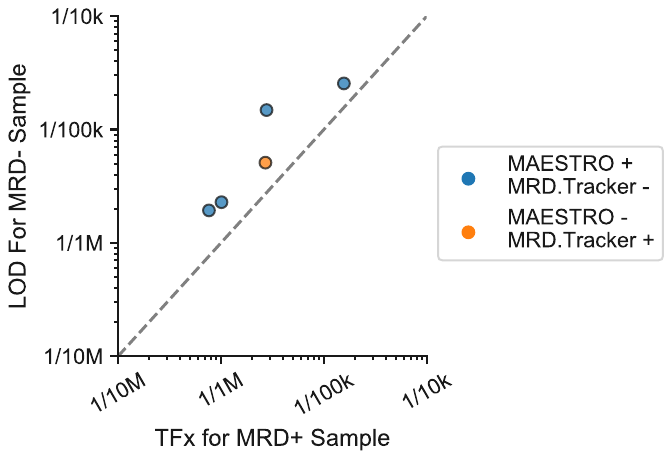

**Supplementary Figure 2: Samples with discordant MRD calls can be explained by their LOD.** For every instance where MAESTRO and MRD.Tracker had discordant results, the ctDNA negative sample had a higher LOD than the observed TFx in the positive sample, suggesting that the sample was underpowered.

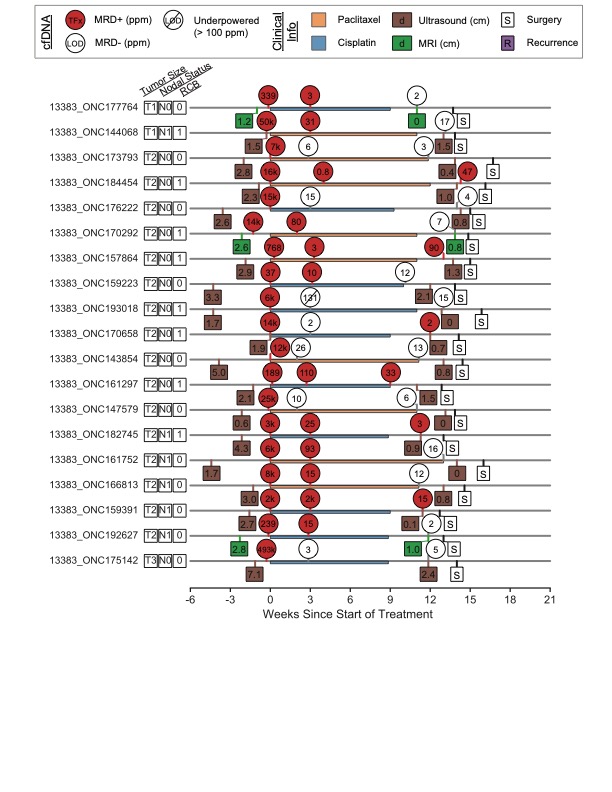

**Supplementary Figure 3: Swimmers plot depicting ctDNA and clinical information for responders (RCB 0/1).** Patients are sorted by tumor size and nodal status and are case-control matched line-by-line with Supplementary Fig. 4.

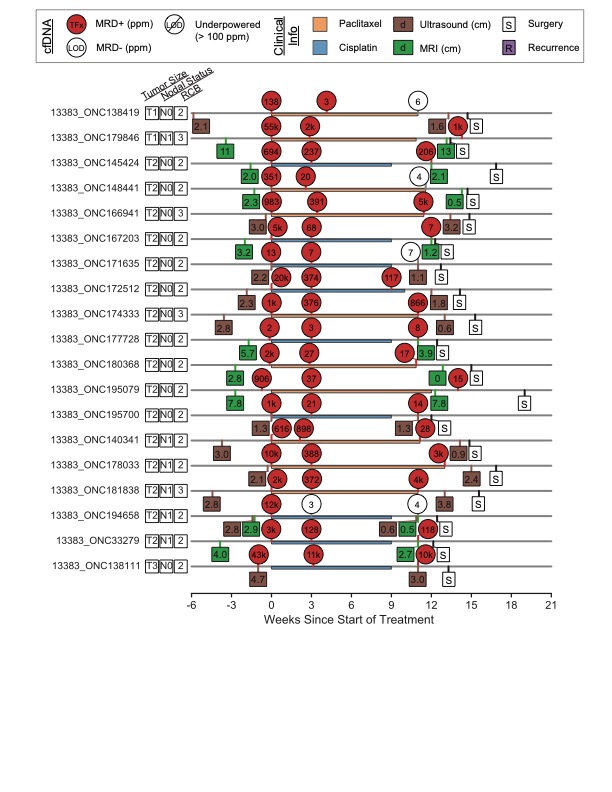

**Supplementary Figure 4: Swimmers plot depicting ctDNA and clinical information for non-responders (RCB 2/3).** Patients are sorted by tumor size and nodal status and are case-control matched line-by-line with Supplementary Fig. 3.

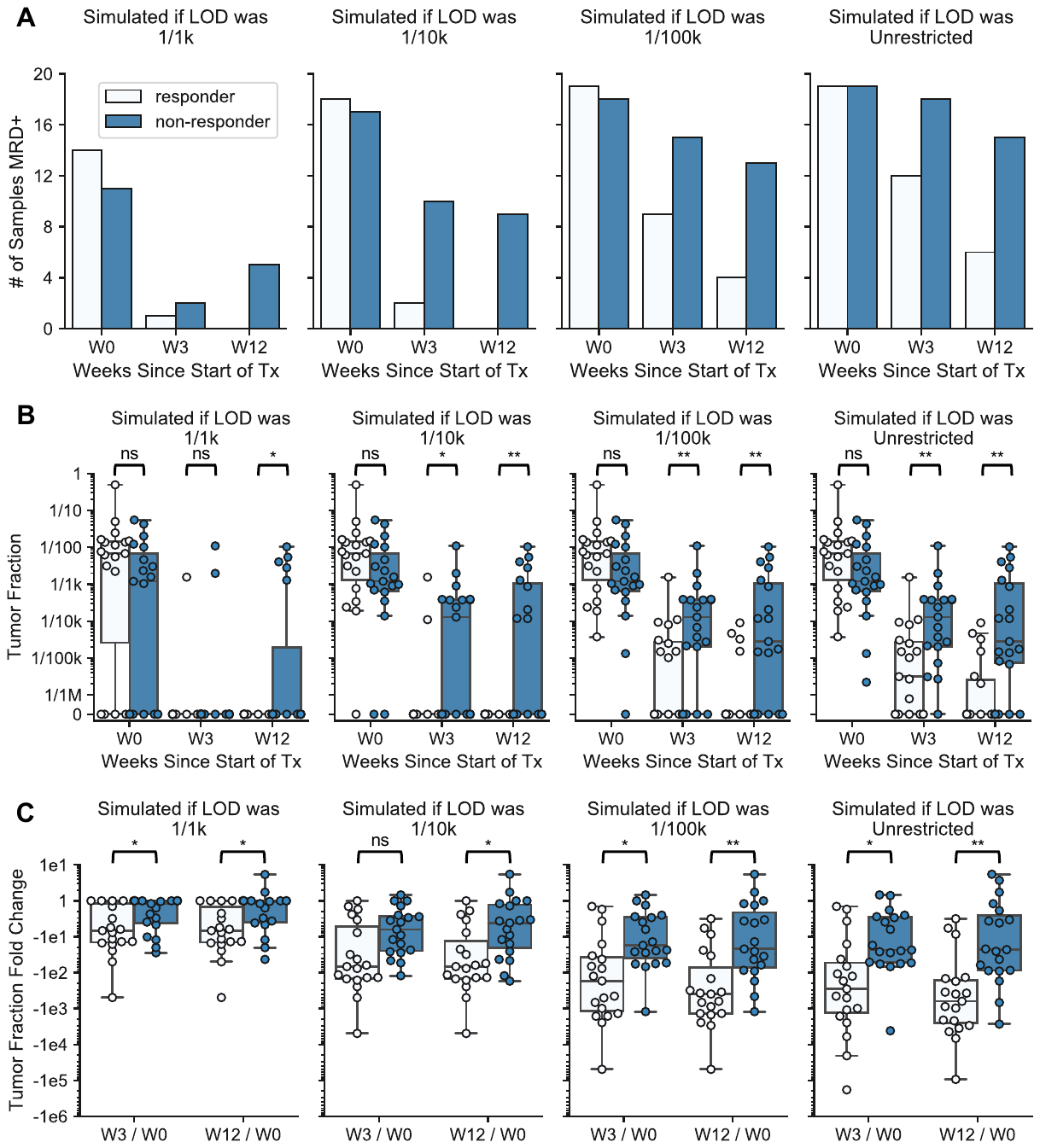

**Supplementary Figure 5: Simulating ctDNA results if LOD was comparable to existing tests.** We simulated sensitivity thresholds to understand its impact on MRD calling and response prediction. At the various thresholds, TFx estimates below the threshold were assumed to be 0. The panels above depict the resulting changes in (A) the number of samples called MRD positive, (B) the TFx distributions and (C) the TFx fold change distributions. Statistical significance was evaluated using the Wilcoxon signed-rank test (*: p < 0.05; **: p < 0.01).
